# PCA-Guided Separation of Mixed Motor Unit Sources in High-Density EMG

**DOI:** 10.64898/2026.06.27.26356748

**Authors:** Zuyu Du, Lara McManus

## Abstract

**Objective:** Decomposition of high-density electromyographic signals enables non-invasive analysis of individual motor unit (MU) behavior, but reliable interpretation of physiological changes in health and disease depends on accurate MU discharge detection. This accuracy is compromised by mixed source estimates, where high amplitude peaks are associated with discharges from more than one MU. We introduce a post-decomposition framework to identify and separate suspected mixed sources using PCA-guided source refinement.

**Method:** For each suspected mixed source, extended and whitened EMG vectors were extracted at source peaks and projected into a low-dimensional PCA subspace. This subspace highlighted MU-specific differences across candidate discharges, including subtle or spatially localized features of the spatiotemporal MUAP profile. Clusters in the PCA subspace were used to initialize source estimates for the constituent MUs. During iterative source refinement, source peak amplitudes were reweighted according to the distance of their corresponding points from the associated cluster center. Particle swarm optimization selected the reweighting factor that minimized the coefficient of variation of inter-spike intervals (CoV_ISI_).

**Results:** The algorithm separated mixed MU sources in simulated and experimental HDsEMG data. In simulated data, resolving mixed sources increased median rate of agreement (RoA) by >40%. In experimental recordings, MU yield increased by 1.27 per trial and CoV_ISI_ decreased by 0.28 (33% RoA improvement).

**Conclusions:** PCA-based representation enhanced separability between MUs with similar MUAP profiles, while distance-based amplitude reweighting reduced re-merging during source refinement. Significance: This framework resolves merged MU discharge trains, improving decomposition accuracy and recovering MUs that might otherwise be excluded by quality thresholds.

## I. Introduction

Decomposition of high-density surface electromyographic (HDsEMG) signals provides non-invasive access to individual motor unit (MU) firing times and action potential properties. This MU level information makes it possible to examine how MUs are recruited, how their discharge rate and timing change, and how these patterns vary across tasks, conditions or populations. This makes HDsEMG a powerful tool for investigating muscle physiology [1], motor control [2], fatigue [3], aging [4], prosthetic control [5], and neuromuscular disorders [6]. However, reliable interpretation of physiological or disease-related differences depends on the accurate detection of MU discharges.

One factor that can comprise the decomposition accuracy is the extraction of mixed source estimates, in which a single extracted source contains discharges from more than one MU [7, 8]. In practice, these mixed MU sources are commonly identified during manual inspection of the candidate discharge trains from the decomposition output [9]. These trains may exhibit characteristics associated with less reliable MU discharge identification. This includes low silhouette index (SIL), low pulse-to-noise ratio (PNR), and irregular discharge timing, reflected by a high coefficient of variation of the inter-spike intervals (CoV_ISI_), although high PNR can still occur for a merged discharge train when the train is well separated from other active MUs [10]. Importantly, these features are not unique to merged MU discharge trains, making it difficult to distinguish a low-quality MU source output from a mixed MU source during manual inspection. This distinction matters because a candidate discharge train dominated by one MU may be salvageable through manual editing, whereas a mixed source with no discernible separation between constituent MUs is unlikely to be recoverable. Current manual editing procedures therefore typically reject these suspected merged MU trains from further analysis, but in some cases they can be resolved by subtracting already identified MUs or by manually separating groups of spikes when the constituent units have clearly heterogeneous spike amplitudes in the source signal [8, 11]. Lundsberg et al. [11] proposed a related automated strategy for separating mixed MU sources, in which short segments extracted around detected peaks in a single source output were clustered. When distinct clusters were detected, this strategy allowed constituent MU discharge trains to be resolved. However, it may not always be possible to identify distinct clusters in this way, particularly in cases of high MUAP similarity, where peaks of similar amplitude may occur due to discharges from more than one MU.

Mixed MU sources are more likely to occur when the constituent MUs have similar spatiotemporal MU action potential (MUAP) profiles across the electrode grid. In these cases, the differences that distinguish them may be spatially localized and have only limited influence on the overall estimation of the separation vector. When all channels are equally considered, localized differences may be overshadowed by shared dominant components in the spatiotemporal MUAP profiles. This means that the separation vector estimated for the mixed source may produce similar amplitude peak for discharges from more than one distinct MU when applied to the extended, whitened EMG. The most challenging mixed sources to identify and separate are these cases in which MUAP similarity means that constituent MUs do not form clearly separable peak groups in the source signal, either by inspection or clustering. To the best of our knowledge, there is no existing framework to identify and separate these challenging mixed source estimates.

To address this gap, this study proposes a post-decomposition framework that first uses clustering in a principal component analysis (PCA) subspace to identify constituent MUs in suspected mixed sources and then separates their discharge trains through subspace-guided reweighting of source peaks. Applying PCA to extended, whitened EMG vectors extracted at larger amplitude peaks in a mixed source can reveal systematic differences across candidate discharges caused by subtle or spatially localized variations in the underlying MUAPs. The resulting PCA subspace emphasizes the parts of the spatiotemporal MUAP profiles that are most informative for distinguishing constituent MUs, allowing their corresponding points to separate into distinct clusters. These clusters are used to initialize new source estimates, which are iteratively refined using distance-based reweighting to prevent convergence back to the mixed source. This approach improved decomposition accuracy and MU yield in simulated HDsEMG, and recovered additional, more physiologically regular discharge trains in experimental recordings. By recovering MUs that would otherwise remain unresolved in the decomposition output or be rejected during editing, this framework improves both MU yield and discharge identification accuracy. This could potentially expand the usable information extracted from HDsEMG decomposition and enable a more reliable characterization of MU behavior.

## II. Theory and Algorithm

This section describes the theoretical framework and algorithm underlying the proposed post-decomposition method for resolving mixed MU sources. In Section II-A, the HDsEMG signal model and source estimation procedure are introduced to establish notation for the extended and whitened EMG representation, separation vectors, and source outputs. Section II-B describes the mixed MU source, which is the problem addressed in this study. Section II-C first outlines the PCA-based representation used to examine discharges detected within suspected mixed sources, then details the clustering procedure used to identify candidate constituent motor units. Section II-D introduces the procedure for unmerging mixed sources, specifically the subspace-guided reweighting and iterative refinement steps used to recover separate discharge trains. Section III-A and III-B describe the generation of the simulated and experimental data used in this study, and section III-C presents the evaluation metrics used to access the performance of proposed algorithm.

### A. Signal model and source estimation

HDsEMG signals can be modelled as a convolutive mixture of multiple MU discharge trains and their corresponding MUAPs, plus additive noise:

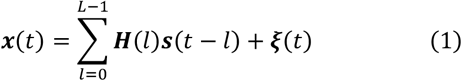

where ***x***(*t*) ∈ ℝ^M×1^ is the vector of HDsEMG signals recorded from *M* electrodes at time *t*, ***s***(*t*) ∈ ℝ ^N×1^ is the vector of binary discharge signal of *N* activated MUs and ***ξ***(*t*) ∈ ℝ^M×1^ represents additive noise. The matrix ***H***(*l*) ∈ ℝ^M×N^ is the lag-dependent mixing matrix in which each element *h*_*mi*_ (*l*) represents the contribution of the *i*-th MUAP to the *m*-th HDsEMG channel at lag *l*. L denotes the duration of the motor unit action potential in samples.

The original convolutive mixing model can be represented as an equivalent linear mixture model by extending the HDsEMG signal to include *R*-1 delayed versions of each channel, where *R* is the extension factor:

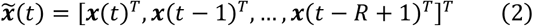

with

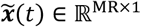

This gives the extended linear mixture:

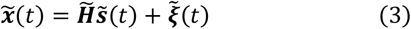

where 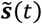 represents the extended source signal and 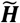 is the corresponding extended mixing matrix, which contains the lagged MUAP contributions across channels, for different time delays. This extended representation incorporates the temporal information contained in the MUAP waveforms and provides the linear mixture form required for signal whitening and subsequent blind source separation [12].

To reduce second-order correlations between components of the extended signal and normalize its variance, the extended HDsEMG signals were whitened by applying a linear transformation:

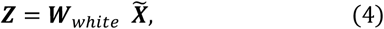

where

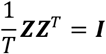

Here, 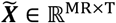 is the matrix of extended HDsEMG signals:

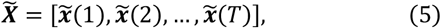

and ***Z*** ∈ ℝ^MR×T^ is the whitened, extended HDsEMG signal matrix:

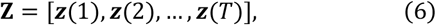

where ***z***(*t*) ∈ ℝ^MR×1^ is the whitened, extended HDsEMG signal at time t, here referred to as an observation vector. ***W***_*whiten*_ ∈ ℝ^MR×MR^ is the whitening matrix, and *T* is the signal length in samples.

Based on the extended linear mixture model, MU sources components can be iteratively estimated from the whitened signals using FastICA [13]. In each iteration, a normalized separation vector ***ω***_i_ ∈ ℝ^MR×1^ was optimised and used to estimate *i*-th source component, ***y***_*i*_:

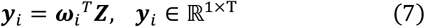

Candidate discharge times were identified from ***y***_*i*_ using of peak detection and k-means++ clustering [14]. The separation vector was then refined using convolution kernel compensation (CKC) [12, 15] by averaging the observation vectors at the candidate discharge detected within the same estimated source component:

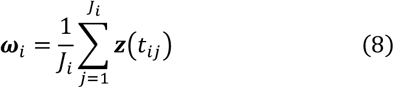

Here, ***z***(*t*_*ij*_) ∈ ℝ^MR×1^ is the observation vector at the *j*-th candidate discharge time, *t*_*ij*_, detected from the *i*-th estimated source component, where ***J***_*i*_ is the total number of candidate discharges detected within that component. After decomposition, only sources that met the predefined quality criteria were retained, and duplicate sources introduced by signal extension were removed.

### B. Mixed MU source problem

A mixed MU source is a possible output of HDsEMG decomposition, in which a single extracted source contains candidate discharges originating from more than one MU. These sources are typically characterized by an irregular or physiologically implausible discharge pattern. Although many low quality source outputs are rejected using typical acceptance quality criteria, mixed sources may still exhibit acceptable SIL or PNR values when the discharge events are well separated from background activity. Mixed sources can reduce the final MU yield when excluded, while those that pass quality criteria may bias subsequent analyses if retained. Therefore, mixed MU sources must be identified and either separated into their constituent MU discharge trains or excluded when reliable separation is not possible.

### C. PCA-based representation and clustering of discharge events

#### 1) Principal Component Analysis-based Representation

For each suspected mixed source, the source signal was obtained by applying the corresponding separation vector to the whitened extended HDsEMG signal, as shown in (7). The amplitude of each candidate discharge was then defined as the value of this source signal at the candidate discharge time.

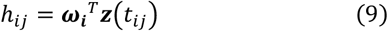

where *h*_*ij*_ denotes the amplitude of *j*-th candidate discharge event in the *i*-th source, ***ω***_***i***_ is the separation vector for that source, and ***z***(*t*_*ij*_) is the observation vector at time *t*_*ij*_.

This scalar amplitude reflects how strongly each observation vector aligns with the separation vector at the time of a discharge event. However, candidate discharges from different MUs may produce similar amplitudes when their MUAPs share dominant spatiotemporal features, even if their waveforms differ in more localized regions of the electrode grid. In addition, observation vectors representing a correctly identified MU firing may also not be perfectly aligned with its separation vector due to noise and MU superposition. This means that it may not be possible to separate constituent MUs in a mixed source based on amplitude alone. The overall post-decomposition framework proposed to identify and separate suspected mixed sources is summarized in Fig. 1.

**Fig. 1:**
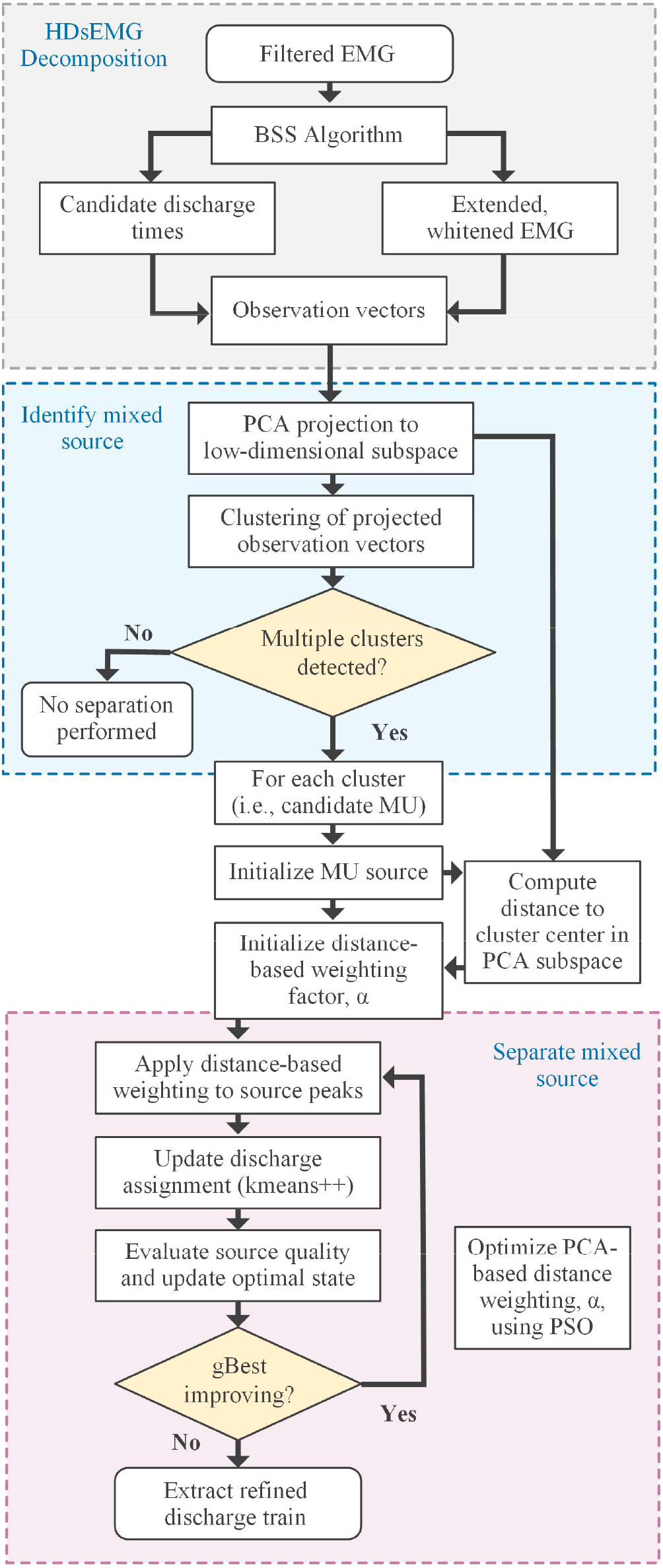
Flowchart of the proposed post-decomposition framework for separating suspected mixed MU sources. The extended, whitened HDsEMG signals are decomposed using BSS to obtain candidate discharge events. These events are used to extract the observation vectors representing the spatiotemporal waveform at each candidate discharge time. These observation vectors are then projected into the PCA subspace and clustered. If multiple clusters are identified, the discharge events corresponding to each cluster are used to initialize separate candidate MU source estimates. Source reconstruction is then refined using distance-based reweighting of source peaks, with the contribution of PCA-subspace distance optimized using particle swarm optimization. The refined discharge train are extracted when the updated source met the required quality criteria.

Although source amplitude provides a useful one-dimensional summary, it reflects only the component of the observation vectors captured by the separation vector. Information contained in other components may contain additional distinguishing features, particularly when constituent MUs have similar dominant spatiotemporal MUAP profiles. PCA was therefore applied to observation vectors extracted at the candidate discharge times of each suspected mixed source to summarize the main differences between candidate events in a low-dimensional subspace. The observation matrix for the *i*-th source was constructed as follows:

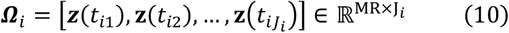

where each column represents the observation vector at a candidate discharge event. Each observation vector, ***z***(*t*_*ij*_), represents the spatiotemporal waveform detected at each event. The mean observation vector was then calculated as:

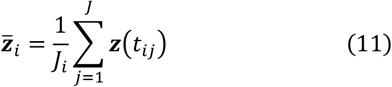

and subtracted from each column of ***Ω***_*i*_ to obtain the mean-centered observation matrix:

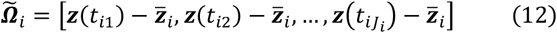

The covariance matrix was then computed as:

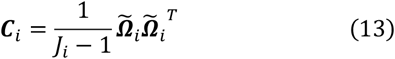

Eigenvalue decomposition is then performed on the covariance matrix [16]:

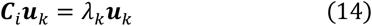

where λ_*k*_ and ***u***_*k*_ denotes the k-th eigenvalue and eigenvectors, respectively. The eigenvectors corresponding to the two largest eigenvalues were retained to form the PCA subspace:

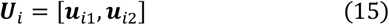

The mean-centered observation matrix was then projected onto this subspace:

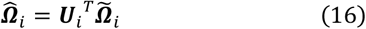

where 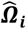 contains the first two principal component scores for the candidate discharge events of the of *i*-th source.

The PCA scores then summarize the main patterns of variation across observation vectors in a low-dimensional subspace. Because the principal component directions are determined by the features that vary most across candidate events, this representation can emphasize subtle or localized features of the spatiotemporal MUAP profile that contribute most strongly to differences between similar MUs. This concept is illustrated in Fig. 2, where localized differences between similar MUAP profiles that are not apparent from the original source amplitudes become more distinguishable after PCA-based representation. Candidate discharges were then clustered in the PCA subspace to determine whether the mixed source contained separable groups corresponding to distinct MUs.

**Fig. 2:**
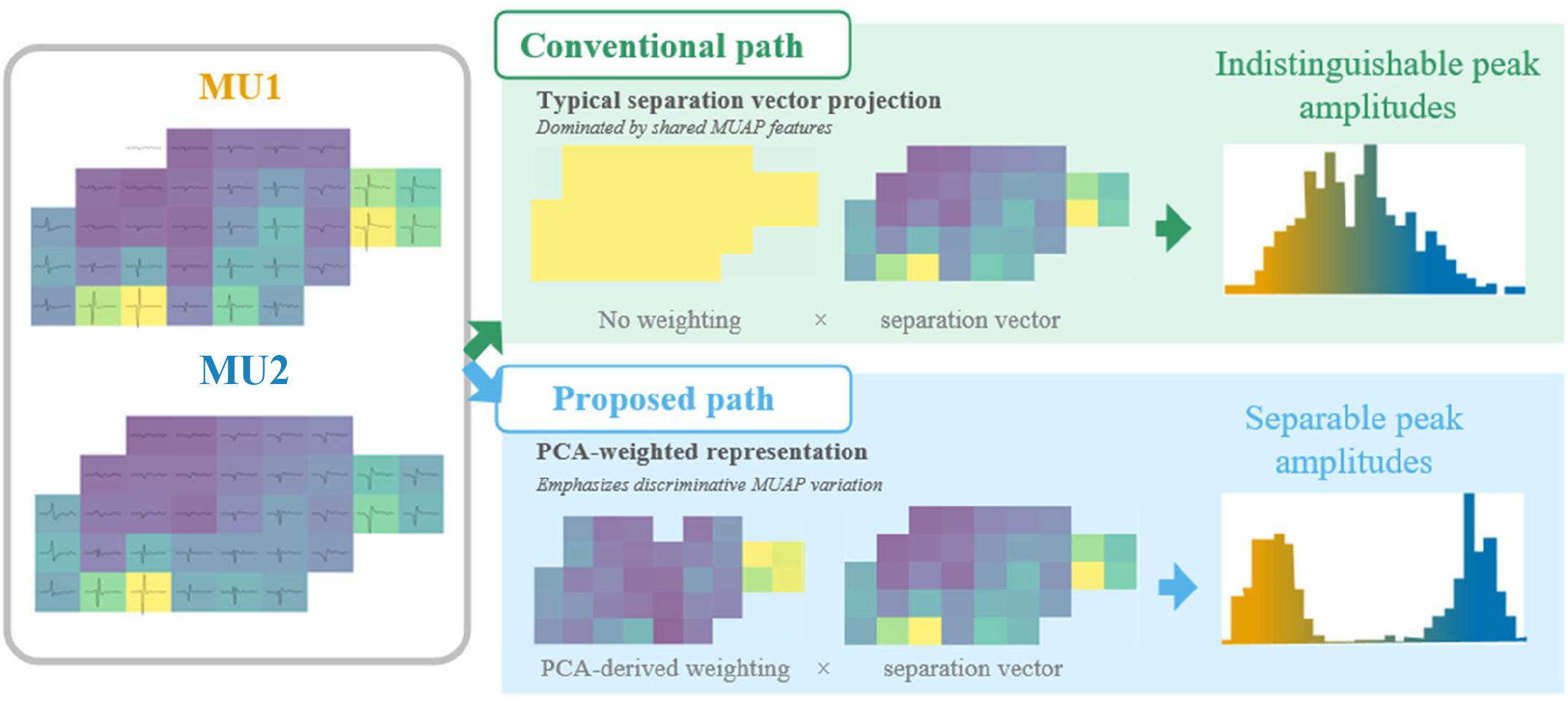
Illustration of PCA-based separation of MUs with similar MUAP morphologies. When MUAPs from different MUs share dominant features, their candidate discharges may produce similar source amplitudes and appear as a mixed source. Subtle differences that are localized to specific channels may be masked in the one-dimensional source signal. PCA represents each candidate discharge using weighted combinations of the spatiotemporal observation-vector components, giving greater influence to features/channels that explain variation between candidate events. This can make discharges from different MUs more separable in the PCA subspace. The PCA-derived separation is subsequently used to guide distance-based reweighting of source peaks during iterative reconstruction., see Section II-C.

#### 2) Clustering of candidate MUs

As MU source estimates with SIL values above 0.95 were considered unlikely to represent mixed sources [17], only source estimates with SIL below 0.95 were included in the proposed separation procedure. A representative example of a suspected mixed source is shown in Fig. 3. In this example, discharges from three distinct MUs produced high-amplitude peaks in the source signal, which could not be distinguished based on peak amplitude alone. However, representing the corresponding observation vectors in the PCA subspace revealed three discernible clusters. This illustrates how the PCA-based representation of observation vectors can reveal differences in MUAP morphology that are not readily apparent from the one-dimensional source signal, providing a basis for identifying mixed sources.

**Fig. 3:**
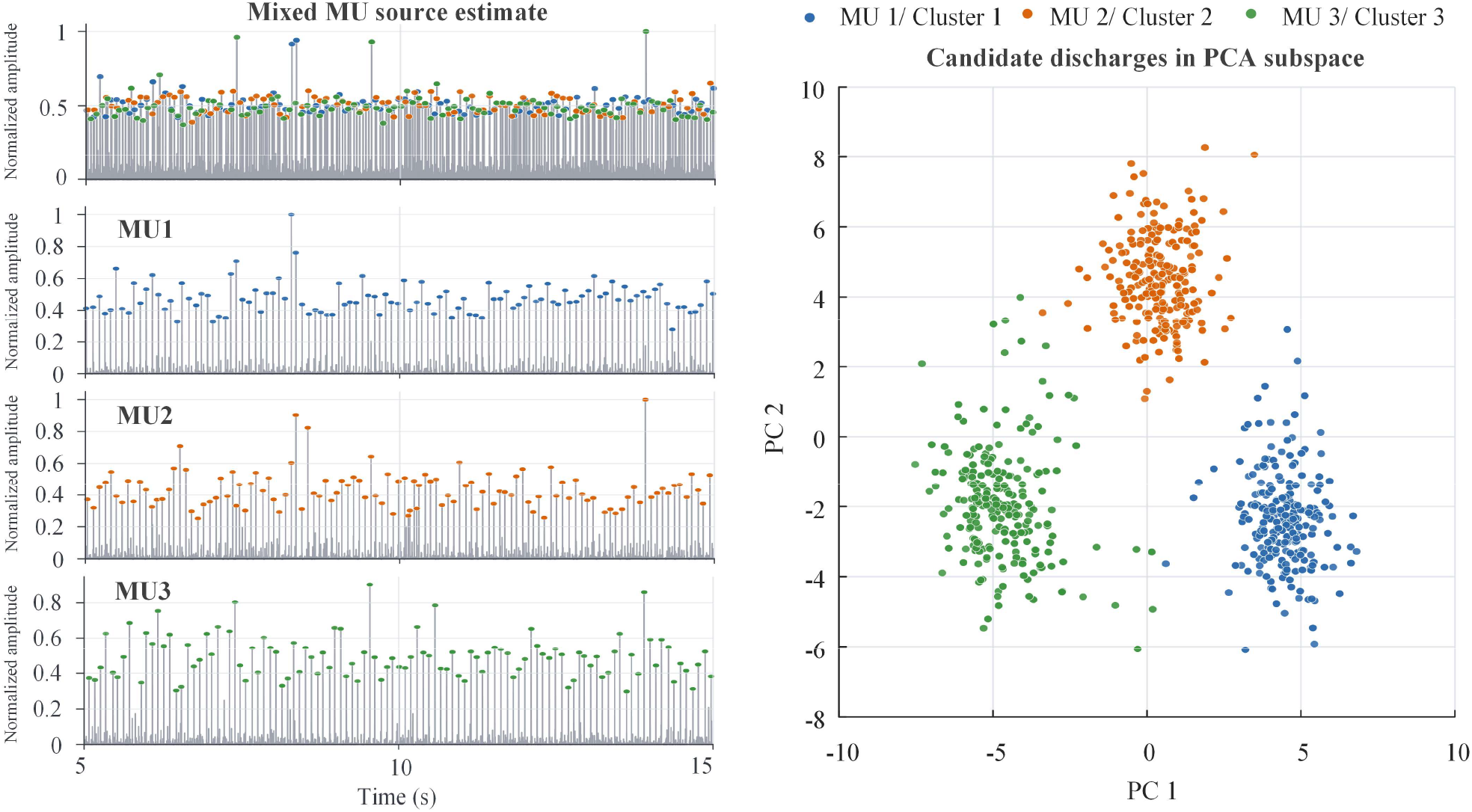
PCA-based clustering of a suspected mixed MU source. Candidate discharge events from a mixed source form three distinct clusters in the PCA subspace. Each cluster represents a candidate MU and can be subsequently used to initialize a separated MU source.

Visual inspection of the PCA subspace can be used to assess whether candidate discharges formed separable groups, as shown in Fig. 3. However, to reduce the need for manual assessment and provide an automated estimate of cluster presence and number, candidate discharge events were also clustered using a mean-shift algorithm [18]. Mean-shift is a density-based clustering algorithm that groups points according to regions of high local point density, allowing clusters to be identified without specifying the number of clusters in advance. This approach does not assume a predefined cluster shape, is relatively robust to outliers, and is not sensitive to initialization. The bandwidth parameter, which controls the neighborhood of density estimation, was selected empirically. To ensure sufficient separability between identified clusters, pairwise cluster separability was assessed using the silhouette coefficient. Cluster pairs with a silhouette value below 0.75 were merged and clusters containing less than 50 points were excluded from further analysis. For the experimental data, cluster assignments were also visually inspected to confirm whether the identified groups represented distinct and separable clusters in the PCA subspace, with assignments manually corrected where necessary. Subsequent processing was performed using both the automatically generated clusters and the visually inspected cluster assignments to evaluate the effect of manual inspection on the separation procedure. Each retained cluster was treated as a candidate MU source for subsequent reconstruction. Sources with fewer than two retained clusters were not considered mixed sources for the purpose of the proposed separation procedure.

### D. Separating Mixed MU Sources

#### 1) PCA-guided peak reweighting

Each retained PCA cluster provided the initial set of discharges for a new candidate MU source. For each identified cluster, an initial separation vector was computed by averaging the observation vectors assigned to that cluster, as described in (8). This separation vector was then applied to the whitened, extended HDsEMG signal to obtain an initial source estimate. Because the initial cluster may not contain all the discharges of the MU it represents, the source was iteratively refined using the CKC procedure described in [12] to recover the additional discharge instances. In this process, peaks in the source estimate are detected and clustered into low and high-amplitude groups using k-means++ [19]. The observation vectors corresponding to the high-amplitude peak group are then averaged to update the separation vector. The updated separation vector is reapplied to the whitened extended HDsEMG signal to re-estimate the source, and this process is repeated until convergence.

However, when several MUs have similar spatiotemporal MUAP profiles, candidate discharges from non-target MUs may also produce high-amplitude peaks in the reconstructed source. These peaks can be incorporated into the high-amplitude group during subsequent iterations, causing the source estimate to converge towards a different MU or revert back to the original mixed source. This can occur even when all the discharge events within the initial cluster are correctly identified for the target MU. An example of this re-merging process is shown in Fig. 4.

**Fig. 4:**
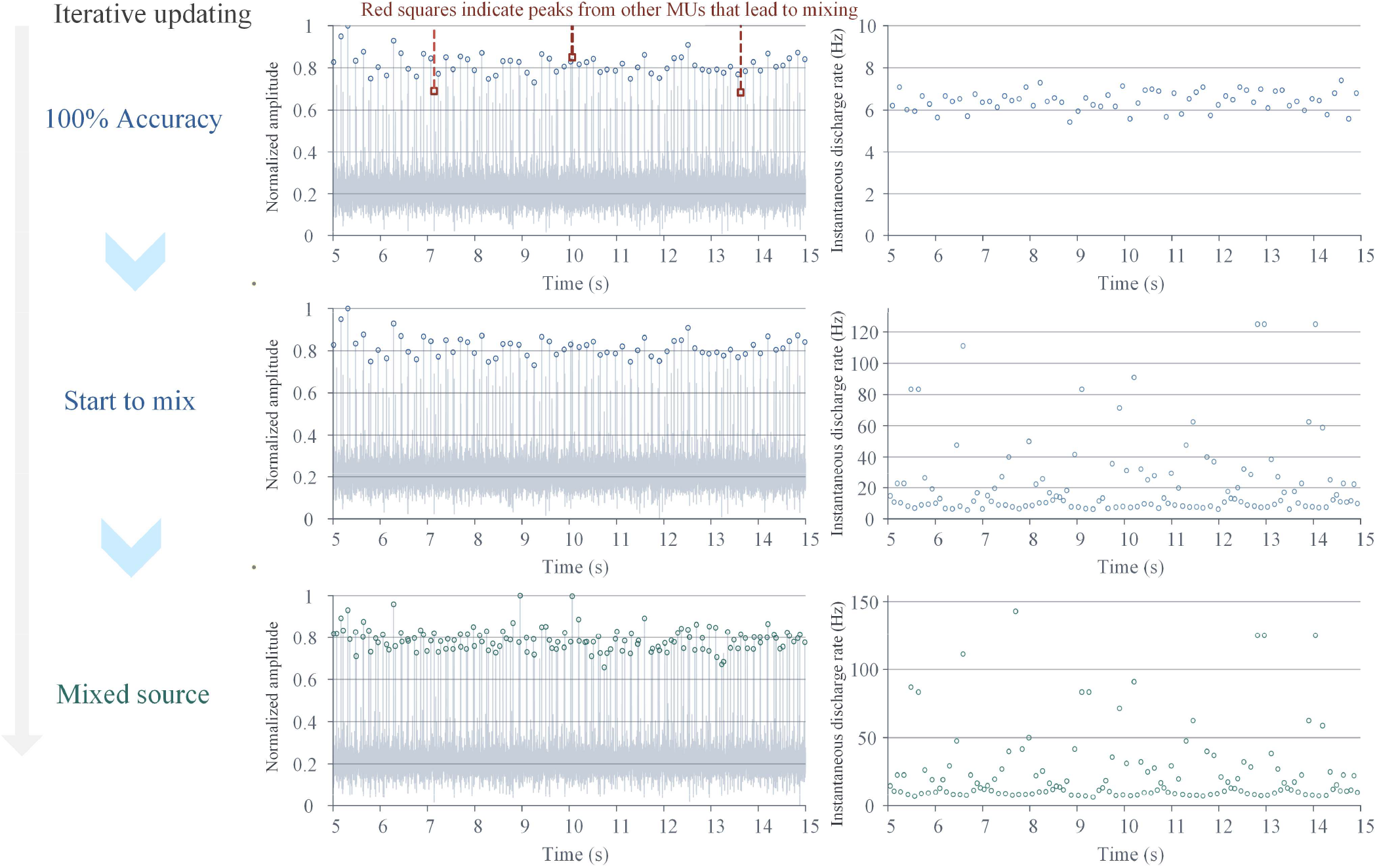
Illustration of the re-mixing process during iterative source refinement. At the initial stage, the simulated source estimate contains accurately identified discharges from the target MU (100% accuracy). However, large amplitude peaks from a non-target MU are also present in the source signal. During iterative refinement, these non-target peaks may be included in the high-amplitude group used to update the separation vector, producing abnormally high instantaneous discharges. As additional non-target discharges are incorporated across iterations, the source estimate can converge back to a mixed source.

To reduce the likelihood that non-target discharges were incorporated during iterative refinement, a subspace-guided reweighting step was applied to the peak amplitudes of the candidate discharges before selecting the high-amplitude group. In the PCA subspace, as illustrated in Fig. 3, candidate discharges belonging to different MUs occupy distinct regions. Therefore, the distance between a point and the target cluster center was used to assess the likelihood that the discharge associated with that peak belonged to the target MU. This was based on the assumption that discharges from the target MU would produce observation vectors with similar spatiotemporal features to those used to define the cluster and would therefore lie close to the cluster center in the PCA subspace. Peaks within a suspected mixed source were thus weighted according to the distance of their corresponding point from the target cluster center. This meant that peaks close to the target cluster were largely preserved, whereas peaks further from the cluster center were penalized before the next refinement step.

After the initial source estimate was obtained from the target cluster, the discharges identified were iteratively refined. At each iteration, peaks were detected in the current source estimate. For each candidate peak, the corresponding observation vector was projected into the PCA subspace defined for the suspected mixed source:

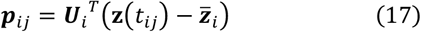

where ***p***_*ij*_ is the PCA score vector for the *j*-th candidate peak valuated during refinement of cluster *c* from the *i*-th suspected mixed source. ***U***_*i*_ contains the first two principal component directions and 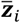 is the mean observation vector defined previously.

For a target cluster *c*, the Euclidean distance between each point and the cluster center was then calculated as:

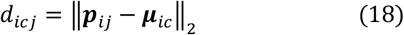

where ***µ***_*ic*_ is the center of cluster *c* in the PCA subspace. The cluster center and distances were defined from the original PCA clustering of the suspected mixed source and was not updated during source refinement. For each iteration, distances were normalized across all candidate peaks detected in the source estimate reconstructed from that cluster. The original peak amplitude was then reweighted as:

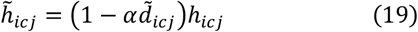

where *h*_*icj*_ and 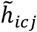 represent the original and reweighted amplitudes of the *j*-th candidate peak, and *α* is the reweighting factor controlling the influence of PCA-subspace distance. When *α* is equal to 0, no distance-based penalty is applied and the procedure is equivalent to the conventional iterative procedure based on peak amplitude. Larger values of *α* increase the penalty applied to peaks corresponding to points further from the target cluster center. The value of *α* was optimised using particle swarm optimisation, as described in Section D-2.

The reweighted amplitudes, 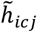 then replace the original peak amplitudes before k-means++ separation into low and high groups. The remainder of the refinement followed the conventional CKC procedure described above, with the high-amplitude group used to update the separation vector until convergence. The final converged source estimate was retained as a candidate separated MU source.

#### 2) Particle Swarm Optimization

The reweighting factor *α* controls the influence of PCA subspace distance on candidate peak amplitudes and thus needs to be optimized. To do this, a metric was required to evaluate the discharge trains obtained using different values of *α*. The coefficient of variation (CoV) of inter-spike intervals (ISI) is commonly used to assess discharge regularity [12]:

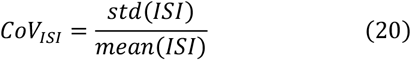

For given MU, a better source estimation typically yields a more physiologically plausible discharge train with more regular inter-spike intervals and therefore a lower CoV_ISI_. However, as CoV_ISI_ is not guaranteed to vary smoothly or differentiably with *α*, a gradient-free optimization method was used. Particle swarm optimization (PSO) is a population-based stochastic optimization algorithm [20]. It utilizes a population of particles to iteratively search for an optimal solution based on a predefined evaluation metric, in this case, CoV_ISI_. PSO has also been recently utilized during the initial BSS stage in HDsEMG decomposition to select source-specific contrast functions [21]. In this study, each particle represented a candidate value of the reweighting factor *α*. At each iteration, the position of the particle was updated according to its previous velocity, personal best solution and the global best solution across all particles:

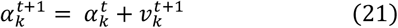

where 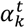 is the candidate value of the reweighting factor represented by the *k*-th particle at iteration *t*, and 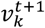 is its updated velocity. The particle velocity is updated by:

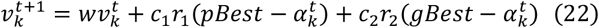

where *pBest* is the best value of *α* previously identified by the *k*-th particle and *gBest* is the best value identified across all particles. The inertia coefficient *w*, cognitive coefficient *c*_1_, and social coefficient *c*_2_ are hyperparameters that jointly regulate the particle updating process by balancing the contributions of its current trajectory, personal best, and global best. The *r*_1_ and *r*_2_ are samples of a z-scored normal distribution.

For each candidate value of *α*, the source refinement procedure described in Section D-1 was performed using the corresponding distance-reweighted peak amplitudes and the resulting discharge train was evaluated using CoV. To reduce the risk of optimizing *α* towards a discharge train corresponding to a different MU, a penalty of 1 was added to the CoV_ISI_ when the estimated discharge train had a low matching rate with the initial discharge train. The penalized CoV_ISI_ was therefore used as the objective function for PSO.

In this study, 10 particles were used. The inertia coefficient was set to *w* = 0.7, and the cognitive and social coefficients were set to *c*_1_ = *c*_2_ = 2. The particles were iteratively updated until convergence, and the final *gBest* value was selected as the optimal reweighting factor. This value was then substituted into (19) to compute the distance-reweighted peak amplitudes for the final MU source estimate.

#### 3) Duplicates Removal

Duplicates MUs can occur among the separated MUs from different mixed sources. To detect duplicate discharge trains, the matching rate of each pair of MUs was computed as:

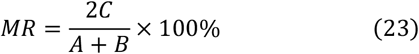

where *MR* is the matching rate, *C* is the number of common discharge occurring within ±10 ms, and *A* and *B* are the total numbers of discharges in the two MUs, respectively. The MU pairs with MR > 30% were considered duplicates, as recommended in [22]. For each duplicate pair, the MU with the higher CoV_ISI_ was removed. Some of the MUs that are recovered during the separation of mixed MU sources may also be duplicates of already identified MUs. Duplicate detection was performed again after separation, comparing the newly recovered MUs to the initial decomposition output before calculating the increase in MU yield. MUs with a matching rate greater than 30% were excluded when quantifying the increase in MU yield.

## III. Materials and Methods

### A. Experimental Datasets

Experimental HDsEMG data from 24 healthy participants (49 ± 19 years old; 10 male, 14 female) were used to evaluate the proposed algorithm. These HDsEMG signals were recorded from the first dorsal interosseous (FDI) muscle during isometric abduction of the right index finger. The study was approved by the St James’s Hospital/Tallaght University Hospital Joint Research Ethics Committee (Project ID: 4831; 2025-Jan-40504050) and conducted in accordance with the Declaration of Helsinki. All participants provided written informed consent prior to participation.

During the experiment, participants sat comfortably with their forearm supported on a custom rest affixed to a heavy table and stabilized using a vacuum cushion and adjustable restraints. The non-index digits and thumb were secured using hook-and-loop straps to minimize force contributions from muscles other than those involved in index finger abduction. The proximal phalanx of the index finger was placed within a ring mount interface attached to a six-degree-of-freedom load cell (MINI40 F/T transducer, ATI Industrial Automation, Apex, USA), which recorded index finger abduction force and off-axis forces. Participants performed isometric index finger abduction while following visual instructions and force feedback presented on a front-facing monitor. Maximal voluntary contraction (MVC) was first determined for each participant as the highest stable force achieved during three short (3 s) maximum contractions, separated by a 3 min rest period. Participants then performed a series of isometric voluntary contractions in which they were required to maintain a constant abduction force for 40 s at either 10%, 20% or 30% MVC.

HDsEMG was recorded from the FDI muscle using a Biosemi ActiveTwo system and a 128-channel electrode grid (FlexPrint, Biosemi, Amsterdam, The Netherlands). The grid consisted of silver/silver-chloride electrode contacts arranged in a 9 × 14 matrix with 5-mm inter-electrode spacing. The grid was positioned over the FDI muscle belly, approximately aligned with the second metacarpal where possible. As the grid size tended to be larger than the FDI muscle, channels judged to be outside the muscle boundary were noted during setup and excluded from analysis. EEG was recorded simultaneously using the same BioSemi ActiveTwo system but was not analyzed in the present study. HDsEMG signals were recorded in monopolar montage at 8192 Hz, with the CMS/DRL electrodes located in the EEG cap. The HDsEMG signals were band-pass filtered between 10 and 500 Hz using a zero-phase 4th order Butterworth filter and down sampled to 2000 Hz. Monopolar and single-differential signals were inspected using root-mean-square (RMS) amplitude and signal-to-noise ratio (SNR), and channels with low SNR, high interference or movement artefact were removed. Single-differential signals were computed offline along the long axis of the 9 × 14 grid. HDsEMG decomposition was performed on the single differential EMG signals and motor unit discharge trains were identified using a blind source separation-based decomposition algorithm [12]. Motor unit action potential templates were estimated by spike-triggered averaging the HDsEMG signals using the detected motor unit discharge times [23].

### B. Simulated Datasets

To construct the simulated HDsEMG signals, motor unit discharge times were generated using a model of the motoneuron pool and muscle force output [24], based on the model described in Lowery and Erim [25]. Motoneurons were simulated as biophysical five-compartment structures consisting of a single soma compartment and four dendritic branches, as described in [26]. Experimentally derived MUAP templates from five participants were used to generate the simulated HDsEMG signals. These MUAPs were obtained using the protocol described in Section III-A. For each participant, the trial with the highest motor unit yield at 10%, 20%, and 30% MVC was first selected. These trials were manually edited, and MUs with a SIL > 0.9 were retained. The remaining MUAPs were concatenated to form five MUAP template sets. Duplicate MUAPs within each set were identified using multi-dimensional template-matching algorithm [27], confirmed by visual inspection, and removed.

Simulated HDsEMG signals were generated by convolving simulated MU discharge trains with the MUAP template sets. Additive Gaussian noise with enhanced low-frequency components was independently added to each channel after low-pass filtering and down sampled to simulate measurement noise. Each HDsEMG dataset consisted of 22.5 s of single-differential HDsEMG with the same spatial configuration and sampling rate as the experimental data. 133 sets of experimentally recorded MUAPs (26.60 ± 3.71 MUAPs for each trial) were used to generate simulated dataset.

### C. Assessment Metrics

For simulated data, algorithm performance was evaluated by quantifying the agreement between estimated and the corresponding ground-truth discharge times. To eliminate the influence of temporal shifts introduced by EMG extension, estimated and reference MU discharge trains were first aligned using cross-correlation, with the maximum allowable shift set to ±10 ms. The rate of agreement (RoA) was then calculated as:

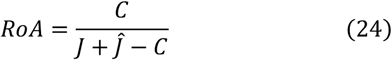

where *C* is the number of estimated discharges occurring within ±0.5 ms of the corresponding ground-truth discharge times, ***J*** and Ĵ denote the total numbers of discharges in the reference and estimated MU discharge trains, respectively.

For experimental data, true discharge times were not known, so the algorithm performance was evaluated by quantifying the reduction in CoV_ISI_ after separation and refinement. Experimental data may contain MUs that do not discharge continuously, resulting in long inter-spike intervals and inflated CoV_ISI_ values. To identify these intermittently active MUs, the proportion of the discharge train duration containing no detected discharges was calculated for each MU. MUs for which these inactive periods exceeded 30% of the total discharge train duration were classified as intermittently active MUs.

To support the use of CoV_ISI_ as a performance metric in experimental data, linear regression was performed on the simulated data to quantify the relationship between CoV_ISI_ reduction and improvements in RoA. Linear mixed-effects models were used to assess whether the final effect of the separation procedure differed across force levels and sex. This was quantified using CoV_ISI_ reduction and the optimal reweighting factor.

### D. Similarity analysis

To investigate whether MUAP similarity contributed to the formation of mixed MU sources, similarity was quantified between pairs of MUs separated from the same mixed source. For the experimental data with manually inspected clusters, all pairs of MUs separated from the same suspected mixed source were defined as separated MU pairs. For comparison, an equal number of control MU pairs was randomly selected from non-merged MUs within the same trial.

For each MU pair, the Euclidean angle, *θ*, between their separation vectors was computed as:

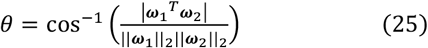

where ***ω***_1_, ***ω***_2_ denote the separation vectors of the two MUs in a given pair. Smaller values of θ indicate greater similarity between separation vectors. To provide a complementary measure of MUAP similarity, the cross-correlation coefficient (CCC) was also computed for each MU pair following the method described in [28]. Two sample t-tests were then used to compare the Euclidean angle and CCC values between separated MU pairs and randomly selected control MU pairs.

## IV. Results

### A. Simulated Datasets

The proposed framework for separating mixed sources was first evaluated using simulated HDsEMG data, where ground-truth discharge times were known. This allowed the relationship between CoV_ISI_ reduction and improvement in RoA to be quantified directly. In the simulated dataset, 38 mixed source estimates (mean: 0.48 per trial; range: 0–3) were identified in the simulated dataset. Separation of these sources produced 81 new MUs after duplicate removal, corresponding to 1.01 separated MUs per trial. No separated MU were duplicates of MUs identified during the original decomposition. The separated MUs showed a median CoV_ISI_ reduction of 0.39 [IQR: 0.29–0.48], and a median RoA improvement of 43% [IQR: 37%–47%], as shown in Fig. 5 (a) and (b). Linear regression showed a strong positive association between CoV_ISI_ reduction and improvements in RoA (β = 0.73, SE = 0.047, F(1,79) = 245, p < 0.001), with CoV_ISI_ reduction explaining 75.6% of the variance in RoA improvement (*R*^2^ = 0.756), as shown in Fig. 5 (c).

**Fig. 5.**
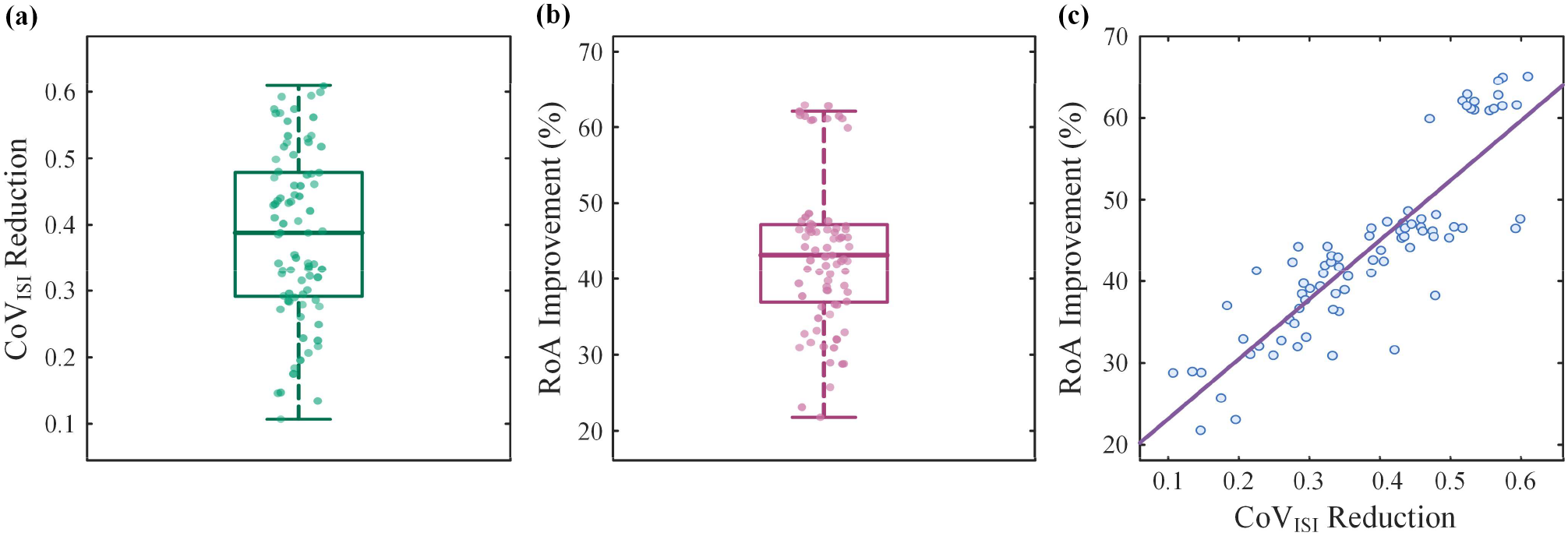
Performance of the proposed separation framework in simulated HDsEMG data. (a) CoV_ISI_ reduction for MUs separated from simulated mixed source estimates. (b) Improvement in rate of agreement (RoA) after separation, calculated using the known ground-truth discharge times. (c) Linear relationship between CoV_ISI_ reduction and DIR improvement in the simulated dataset (*R*^2^ = 0.756). The strong positive association supports the use of CoV_ISI_ reduction as a validation metric in experimental data where the ground-truth discharge times are not known.

### B. Experimental Datasets

The framework was then applied to the experimental dataset, where ground-truth discharge times were not available and performance was therefore assessed using CoV_ISI_ reduction. Across the experimental dataset, a median of 57 [IQR: 48–62] single-differential channels were retained for decomposition after removal of channels with low SNR, movement artefact, or locations outside the muscle boundary. The SIL values of the source estimates subsequently identified as mixed sources ranged from 0.67 to 0.94, indicating that SIL alone is not a consistent indicator of mixed-source contamination.

#### 1) Automatically clustering

Using automatic clustering, 164 suspected mixed sources were identified (mean: 0.56 per trial; range: 0–5). Separation produced 307 MUs after removing duplicates that arose across different mixed sources, this corresponded to 1.04 new MUs per trial. Of these, 37 separated MUs matched MUs already identified in the initial decomposition. In 35 of these cases (94.59%), the separated version had lower CoV_ISI_ than the original decomposition output, indicating improved discharge train quality. After excluding these duplicates, the framework recovered 270 additional MUs, corresponding to 0.92 additional MUs per trial.

Across all separated MUs, the median CoV_ISI_ reduction was 0.22 [IQR: 0.09–0.32; Fig. 6 (a)]. Of these, 120 MUs (39%) were classified as intermittently active. The remaining 187 continuously active MUs (0.64 per trial) showed a median CoV_ISI_ reduction of 0.28 [IQR: 0.18–0.34; Fig. 6 (b)]. Based on the simulation-derived regression model, these CoV_ISI_ reductions corresponded to an estimated median improvement in RoA of 32% for all separated MUs and 36% for continuously active MUs.

**Fig. 6.**
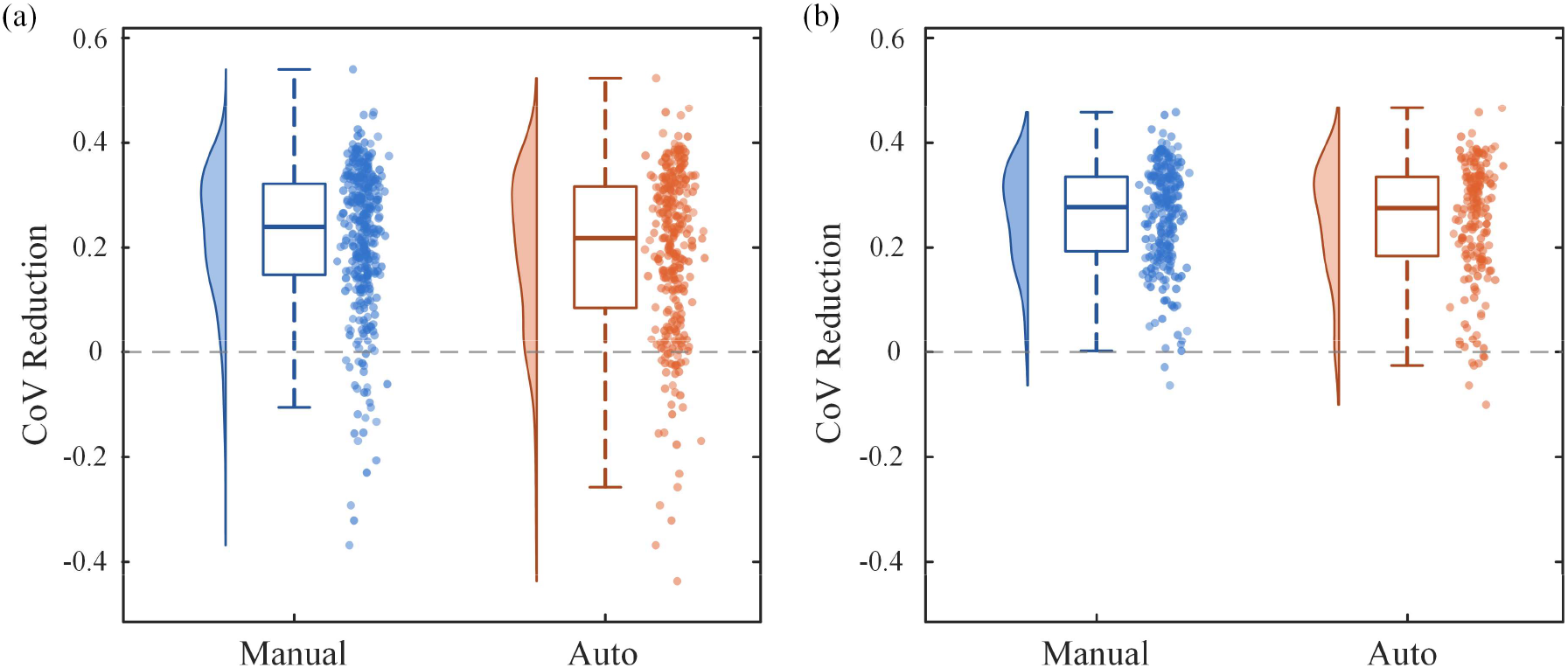
Performance of the proposed separation framework in experimental HDsEMG data. (a) CoVISI reduction of all separated MUs obtained using manually inspected and automatic clustering. (b) CoVISI reduction of separated MUs that are continuously active, after excluding intermittently active MUs, for which long non-discharge periods may inflate CoVISI values.

#### 2) Manually inspected clustering

After manual inspection of the clustering results, 187 suspected mixed MU sources (mean: 0.64 per trial; range: 0–7) were identified. Separation produced 373 MUs after removing duplicates that arose across different mixed sources, this corresponding to 1.27 new separated MUs per trial. Of these, 47 separated MUs matched MUs already identified in the initial decomposition. In 41 of these cases (87.23%), the separated version had lower CoV_ISI_ than the original decomposition output. After excluding these duplicates, the net increase in MU yield was 326, corresponding to 1.11 additional MUs per trial.

Across all separated MUs, the median CoV_ISI_ reduction was 0.24 [IQR: 0.15–0.32, Fig. 6 (a)]. Of these, 122 MUs (33%) were classified as intermittently active MUs. The vast majority of separated MUs that exhibited increases in CoV_ISI_ (i.e. negative values in Fig. 6 (a) and (b)) were intermittently active MUs, where CoV_ISI_ may not fully reflect changes in discharge identification accuracy. The remaining 251 continuously active MUs (0.85 per trial) showed a median CoV_ISI_ reduction of 0.28 [IQR: 0.19–0.34, Fig. 6 (b)]. Based on the simulation-derived regression model, these CoV_ISI_ reductions corresponded to estimated median RoA improvement of 33% for all separated MUs and 36% for continuously active MUs. The optimal reweighting factor was greater than zero in 80.43% of separated MUs, indicating that the reweighting step was an important component of separation. CoV_ISI_ reduction and the optimal reweighting factor did not differ significantly across force levels (p = 0.17) or sex (p = 0.48).

Similarity analysis was performed to determine whether MUs separated from the same mixed source had more similar MUAP profiles than randomly paired MUs from the same trial. A total of 245 separated MU pairs were identified, and an equal number of randomly selected control MU pairs were included for comparison. Separated MU pairs had significantly smaller Euclidean angles between separation vectors than random pairs (t(488) = -31.69, p < 0.001), Fig. 7 (a), indicating greater similarity in their separation vector direction. Separated MU pairs also had significantly higher CCC values (t(488) = 17.97, p < 0.001), as shown in Fig. 7 (b). Although both similarity metrics differed significantly between separated and random MU pairs, the Euclidean angle between separation vectors showed a larger standardized between-group difference than CCC (|d|=2.86 vs. |d|=1.62). This suggests that θ was more sensitive than CCC to differences between distinct MUs with similar MUAP profiles.

**Fig. 7.**
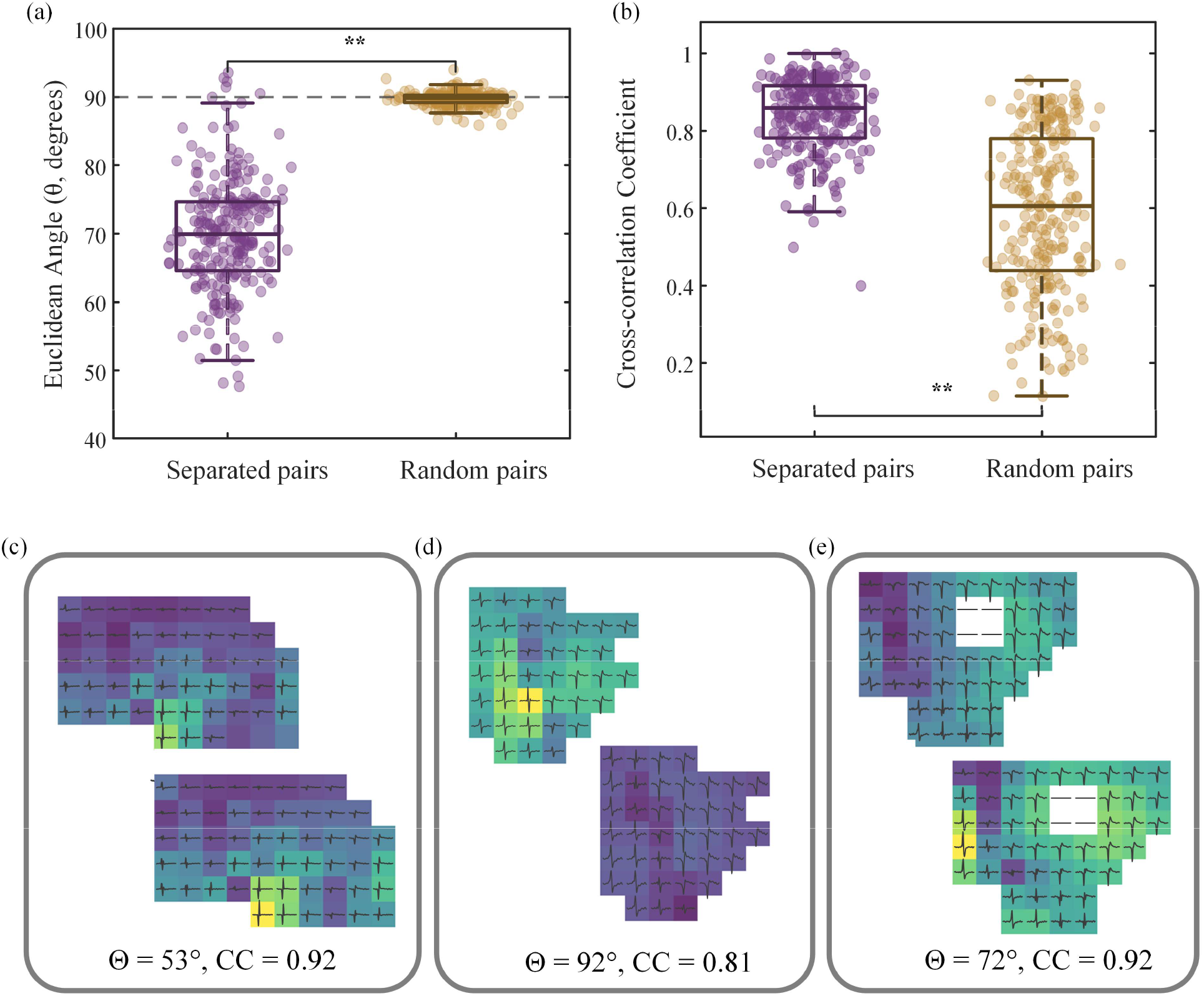
Similarity analysis of separated MU pairs and randomly selected control MU pairs. (a) Euclidean angle, *θ*, between separation vectors for MU pairs separated from the same mixed source and randomly selected control pairs from the same trial. (b) Cross-correlation coefficient between MUAP profiles for separated and random MU pairs. Higher CCC values indicate greater MUAP similarity. (c-e) Representative MUAPs separated MU pairs.

## V. Discussion

This study introduces a novel post-processing framework to separate mixed sources in the HDsEMG decomposition output, where a single extracted source contains discharges from more than one MU. The approach represents the high-dimensional information contained in the extended, whitened EMG observation vectors at each candidate discharge event in a lower dimensional PCA subspace. This reduces reliance on peak amplitude in the one-dimensional source signal and provides a PCA-guided basis for identifying and refining candidate discharge trains from each constituent MU. In simulated data, where ground-truth discharge times were available, the PCA-guided separation framework improved RoA by 43% in the separated MUs, Fig. 5 (b). This corresponded to a 0.39 reduction in the CoV_ISI_ of their inter-spike intervals, Fig. 5 (a). It also recovered an additional 81 MUs from 38 mixed source estimates. The simulation results also supported the use of CoV_ISI_ reduction as a marker of improved discharge identification in experimental HDsEMG data, where ground-truth discharge times could not be directly measured, Fig. 5 (c). In the experimental data, separated discharge trains showed a 0.28 reduction in CoV_ISI_ for continuously active MUs, Fig. 6 (b). This corresponds to an estimated RoA improvement of approximately 36%, based on the simulation-derived relationship shown Fig. 5 (c).

Importantly, the framework also reduces the likelihood that mixed source estimates are retained as contaminated outputs in subsequent analyses. This is relevant because 31% of mixed sources had SIL values above 0.85, a threshold that may be used to accept decomposed MUs in some studies. It may also reduce time spent attempting to manually edit mixed discharge trains that are not readily resolved by amplitude-based inspection of the source peaks.

By improving discharge identification, the framework enables more MUs to satisfy commonly used acceptance thresholds, increasing MU retention and overall yield. Separation increased the MU yield by an average of 1.11±1.71 per trial, although the gain varied considerably across trials and participants according to the prevalence of mixed sources. In trials where mixed sources were present, MU yield increased by an average of 2.89 per trial. The incidence of mixed sources was also highly participant-dependent. In 11 participants mixed sources occurred in over half of all trials, and in these participants, there was a much higher rate of mixed sources (1.2 per trial compared with 0.64 per trial overall). Although it was not assessed in the present study, cluster incidence is also likely to be highly muscle dependent. The complex, bipennate fiber architecture of the FDI muscle [29, 30] may result in more spatially distinct MUAP profiles and a relatively low cluster incidence when compared with other muscles such as the biceps.

The improvement in CoV_ISI_ was slightly lower for intermittently active MUs, Fig. 6 (a), and some of these discharge trains displayed increased CoV_ISI_ after separation. Some MUs may discharge intermittently during the isometric hold, a pattern that can be observed in healthy participants but tends to be more prominent in neurodegenerative conditions like motor neuron disease. Visual inspection of the discharge trains suggested that the increased CoV_ISI_ in these MUs reflected large gaps between periods of MU activity, which inflated ISI variability, rather than a true reduction in discharge identification accuracy. Among continuously active MUs, only two separated MUs showed increased CoV_ISI_ after manual inspection of the clustering results. In these rare cases, the reweighting procedure appeared to be overly conservative – in reducing the inclusion of discharges from the non-target MU, it also excluded a substantial number of discharges from the dominant MU. This resulted in a slight increase in CoV_ISI_ despite successful separation of the mixed source. These outputs can be easily identified and removed during post-processing because they have a high matching rate with the original mixed source but poorer CoV_ISI_ after separation. Together, these observations highlight that CoV_ISI_ is an imperfect proxy for discharge train quality, particularly for intermittently active MUs. The CoV_ISI_ based estimates reported here are therefore likely to underestimate the true performance of the proposed framework.

Mixed sources can arise at difference stages of the HDsEMG decomposition process. During the initial BSS stage, FastICA estimates separation vectors that are intended to isolate the discharges of an individual MUs. However, discharges from distinct MU may produce similar higher order features in the extended, whitened HDsEMG signal. Under these conditions, FastICA could converge to a local optimum in which a single source estimate contains discharges from more than one MU. This is more likely to occur when the corresponding MUAPs have similar spatiotemporal MUAP profiles. In this case, FastICA may estimate a separation vector that captures features shared across MUs rather than more subtle or localized features that could distinguish an individual MU. Some of these mixed sources can be resolved to a single MU during the next CKC refinement stage if the initial separation vector is more strongly aligned with one constituent MU. Discharges from that MU are then more likely to generate higher amplitude peaks in the source signal, enabling the refinement loop to converge towards a single MU source. However, if discharges from more than one MU continue to produce large peaks in the source signal, the refinement procedure may preserve or reinforce the mixed source rather than isolating a single MU. A similar issue can arise during iterative refinement, even when initial source estimate primarily reflects one MU. If discharges from other non-target MUs produce relatively high peaks, they may be increasingly incorporated into the high amplitude peak group identified by k-means++ during iterative refinement. This group is then used to update the separation vector, which can cause the source estimate to converge towards a mixed source. This is a recognized challenge in FastICA-based decomposition, which motivated work to reduce repeated or local convergence using progressive peel-off strategies [31]. Spatiotemporal MUAP similarity is a key factor that increases the likelihood of discharges from non-target MUs being incorporated into the source signal during the decomposition process. This is supported by the findings that pairs of separated MUs have significantly smaller angles between their separation vectors, θ, Fig. 7 (a) and higher cross-correlation values, Fig. 7 (b). However, MUAP similarity was not sufficient to explain all mixed sources. Some separated MU pairs had relatively large separation vector angles and were visually distinct, Fig. 7 (d). This indicates that MUs with more distinct high-dimensional source representations can also become merged. Other factors, such as MUAP superposition and signal quality, are also likely to contribute to the formation of mixed sources. Higher spatial sampling is one approach that may help to reduce the number of mixed sources by capturing more discriminative MUAP features [32, 33].

The Euclidean angle between separation vectors showed a larger standardized between-group difference than CCC and nearly all random pairs occupied approximately orthogonal positions in the high-dimensional space. Together this suggests that θ was more sensitive to differences between distinct MUs with similar MUAP profiles. This was also evident from the CCC distribution, where some separated MU pairs showed very high CCC values (CC > 0.9) despite representing distinct MUs, Fig. 7 (c, e). These findings highlight a limitation of using waveform cross-correlation alone to distinguish unique MUs, or conversely, to track the same MU across trials or interventions based on MUAP shape. Other methods, such as multi-dimensional MUAP matching or these angle-based metrics, may provide a more robust approach for duplicate detection and MU tracking [27, 34].

The proposed framework addresses mixed source estimates in two linked stages. First, it makes it possible to identify mixed sources by incorporating multi-dimensional information from the extended, whitened EMG observation vectors, rather than relying on the one-dimensional source signals. This is important because candidate discharges from different MUs may produce similar peak amplitudes in the source estimate when their MUAPs share dominant spatiotemporal features. Subtle or localized differences between them that are present in their observation vectors may not be apparent in their one-dimensional representation. Representing these observation vectors in a PCA subspace emphasizes the directions of variation that distinguish candidate discharges from different MUs within the mixed source. This can reveal separable groups of discharges within the mixed source, which form clusters in the PCA subspace that can be used to guide reconstruction of each constituent MU.

Once these clusters are identified, they provide a more specific initial discharge set for each constituent MU. Reinitializing the source from a PCA cluster can provide a better starting point for the source estimate, however, it does not prevent discharges from non-target MUs being gradually re-incorporated into the signal during CKC refinement. If non-target MUs still produce high amplitude peaks in the source signal, these peaks can be re-introduced during iterative refinement, causing the source to revert to a mixed output. To reduce this risk, the framework reweights the peaks of candidate discharges according to their distance from the target MU cluster center in PCA space, optimizing α via PSO. This reduces the influence of peaks that are less likely to belong to the target MU and helps the refinement procedure to continue while remaining constrained to the target MU. The fact that the optimal reweighting factor was greater than zero for most separated MUs indicates that this reweighting step was a key element of the separation process.

Despite the irregular cluster shapes and variable separation between constituent MUs in the PCA subspace, the automatic clustering procedure performed relatively well. It identified most separable mixed sources and produced clear improvements in CoV_ISI_ and MU yield without additional manual inspection. However, manual inspection still improved performance by identifying additional MUs that were missed by automatic clustering. These cases often involved clusters that were close together or had irregular boundaries in the PCA subspace, making them difficult to separate using the current mean-shift clustering. Manual inspection was also useful for identifying cases where apparent clusters reflected gradual MUAP shape changes over time rather than distinct MUs. Substantial MUAP waveform changes can occur with fatigue [35] and temperature changes [36], while more subtle changes may also occur under stationary conditions in both healthy and patient populations [37]. Future work could investigate more adaptive clustering approaches, to narrow the performance gap between automatic and manual inspected clustering to make the framework more suitable for fully automated post-processing.

It should be noted that not all mixed source estimates can be successfully separated into their constituent MUs. Source estimates with low SIL often contain discharges from several MUs, but identifying a source as mixed does not necessarily mean that its constituent MUs are recoverable. When too many MUs contribute to the same source estimate it becomes difficult to identify projection directions that selectively separate each MU. The same problem can occur when the recorded MUAP representations vary substantially across discharges of the same MU, particularly when this variation is large relative to the MUAP differences between constituent MUs. Future work could investigate nonlinear approaches to expand this separability limit and recover structure that may not be captured by the current linear PCA representation. Higher signal quality reduces discharge-to-discharge variability in the recorded MUAP representations and helps preserve discriminative MUAP features. Careful skin preparation, electrode placement, and noise reduction therefore remain important for maximizing signal quality and improving separability. It may also be possible to increase MU yield by incorporating PCA-based information earlier in the decomposition process, during the FastICA or CKC refinement stages, building on recent efforts to use MUAP morphology to improve source extraction [38]. Finally, the current framework is best suited to steady isometric contractions, where MUs discharge consistently enough to define stable clusters and support CoV_ISI_-based optimization. Future work could investigate alternative evaluation metrics for dynamic contractions, fatigue, and conditions with intermittent or non-stationary MU firing. This would broaden the applicability of the framework beyond steady isometric contractions and further strengthen its use for studying MU behavior in complex experimental tasks and clinical populations.

## VI. Conclusion

This study introduced a PCA-guided post-processing framework for identifying and separating mixed source estimates in HDsEMG decomposition output. By using the high-dimensional information contained in extended, whitened EMG observation vectors, the framework provides a way to recover constituent MU discharge trains that may not be separable from peak amplitude in the source signal alone. The results show that the MU information contained within mixed sources is recoverable, and separating these merged MU trains can improve both yield and discharge identification. The findings also support MUAP similarity as a key contributor to mixed source formation, while demonstrating that PCA-based representations can reveal discriminative features between constituent MUs. More broadly, this framework provides a step towards more complete and reliable extraction of MU behavior from HDsEMG recordings. It may help to ensure that mixed sources are not discarded unnecessarily or retained as contaminated outputs in subsequent analyses.

## Data Availability

All data produced in the present study are available upon reasonable request to the authors

## Acknowledgment

The authors thank S. Bista, G. Del Duca and B. D’Alpaos for their assistance with the experimental recordings. The authors also thank B. Nasseroleslami for co-supervisory support provided during the PhD research associated with this work.

